# A novel use of diffusion-weighted whole-body magnetic resonance imaging with background body signal suppression to diagnose infectious aortitis

**DOI:** 10.1101/2024.03.11.24304138

**Authors:** Jien Saito, Masahiro Muto, Masafumi Tada, Isao Yokota, Shinji Kamiya, Yukihide Numata, Hideki Sasaki, Takuya Hashizume, Miki Asano, Satoru Wakasa

**Affiliations:** Department of Cardiovascular Surgery, Hokkaido University Graduate School of Medicine, Hokkaido, Japan (JS, SW); Department of Cardiovascular Surgery, Nagoya City University East Medical Center, Aichi, Japan (JS, SK, YN, HS, MA); Department of Biostatics, Hokkaido University Graduate School of Medicine, Hokkaido, Japan (JS, IY); Department of Radiology, Ogaki Municipal Hospital, Gifu, Japan (MM); Department of Health Promotion and Human Behavior, Kyoto University Graduate School of Medicine, School of Public Health, Kyoto, Japan (MT); Department of Neurology, Nagoya City University East Medical Center, Aichi, Japan (MT); Department of Radiology, Nagoya City University East Medical Center, Aichi, Japan (TH)

## Abstract

**Background:** Diffusion-weighted whole-body imaging with background body signal suppression is one of the whole-body magnetic resonance imaging techniques and is effective in diagnosing inflammatory and infectious diseases. We aimed to evaluate the diagnostic performance of this modality in infectious aortitis, which remains unclear.

**Methods:** The study participants were 32 patients with suspected infectious aortitis who underwent computed tomography and magnetic resonance imaging between September 2020 and November 2022. Sensitivity, specificity, and areas under the curve of each imaging modality were studied using a diagnosis based on a combination of imaging results, clinical symptoms, and laboratory tests. Decision curve analysis was performed to determine the benefit of adding magnetic resonance imaging to computed tomography.

**Results:** The median age was 74 years, and 23 participants were men. Fifteen patients (47%) were diagnosed with infectious aortitis. Positive findings for infectious aortitis were identified in 19, 18, and 14 patients by computed tomography, diffusion-weighted whole-body imaging, and the combination of both modalities, respectively. Sensitivity, specificity, and area under the curve for correct diagnosis were 93.3%, 70.6%, and 0.82 (95% confidence interval 0.69–0.95), respectively for computed tomography, 93.3%, 76.5%, and 0.85% (95% confidence interval 0.73%–0.97), respectively for diffusion-weighted imaging, and 86.7%, 94.1%, and 0.90 (95% confidence interval 0.80–0.10), respectively for the combination of both modalities. Decision curve analysis reinforced the clinical benefit of combining the two imaging modalities across all ranges of the probability thresholds.

**Conclusions:** Diffusion-weighted whole-body imaging with background body signal suppression is an effective diagnostic tool for infectious aortitis, especially when combined with computed tomography.

**Clinical Perspective:** Infectious aortitis is a serious disease that is difficult to accurately diagnose. Although PET-CT is associated with high diagnostic performance, limited access to this modality has encouraged the development of an alternative modality. The whole-body MRI with DWIBS is a more available modality, which is commonly used for cancer diagnosis but is also considered effective in diagnosing infectious diseases. The combination of DWIBS and non-contrast CT yielded a sensitivity of 86.7% (95% CI: 59.5–98.3%), a specificity of 94.1% (95% CI: 71.3–99.9%), and an AUC of 0.90 (95% CI: 0.80–0.10) for the diagnosis of infectious aortitis. DWIBS can be a useful modality as an alternative to PET-CT.

## Introduction

Infectious aortitis, such as infectious aortic aneurysm and aortic graft infection, is a life-threatening disease because it is more susceptible to rupture and sepsis.^1^ A high mortality rate was reported in infectious aortic aneurysms, ranging from 26% to 44%.^2^ Therefore, early and appropriate treatment should be performed based on a rapid and reliable diagnosis.

Especially in the case of aortic graft infections, a precise diagnosis is crucial, as these conditions require more invasive treatment.^3^ However, definitively diagnosing infectious aortitis with standard imaging modalities, including computed tomography (CT), is challenging.^4^ Therefore, diagnostic treatment is sometimes administered in cases of suspected conditions.^5^ Positron emission tomography/CT (PET/CT) has emerged as a more precise diagnostic tool for aortic infection.^6–8^ However, PET/CT is not widely used in diagnosing infectious aortitis because it is an off-label use in Japan, is available in limited institutions, and is difficult to perform promptly. Therefore, developing a more feasible imaging modality for infectious aortitis is desired.

Diffusion-weighted whole-body imaging with background body signal suppression (DWIBS) has recently shown promise as an effective diagnostic tool for various conditions, including infectious aortitis. DWIBS was initially developed to evaluate primary tumors and metastases in cancer and has demonstrated diagnostic performance comparable to PET/CT.^9–12^ Its utility in evaluating Takayasu arteritis activity, spine and soft tissue infections, and infectious aortic aneurysms has also been reported.^13–17^ However, no reports evaluated its diagnostic performance for infectious aortitis. Therefore, this study aimed to assess the diagnostic performance of DWIBS for aortic infections in consecutive single-center cases.

## Methods

### Study design

This is a single-center retrospective study. Thirty-five consecutive patients suggestive of infectious aortic aneurysms and aortic graft infections were referred to Nagoya City University East Medical Center between September 2020 and November 2022. Among them, 32 patients who underwent whole-body magnetic resonance imaging (MRI), including DWIBS, within four days before or after the CT scan were studied. Exclusion criteria were if the patient was unable to undergo MRI for any reason or if there was a refusal by the patient to participate in this study. Three patients could not undergo MRI due to shock, inability to remain still, and previously implanted MRI-incompatible stent graft (Zenith; Cook Medical, Bloomington, IN) (Figure 1). Given the absence of consensus on sample sizes for studies evaluating new diagnostic testing methods, the sample size could not be set based on statistical calculations. Instead, we enrolled as many cases as possible, referencing the prior literature.^18–20^ The study protocol was developed and approved by the Ethics Committee of Nagoya City University Hospital (control number: 60-22-0112) before data collection and analysis. This study was also registered at umin.ac.jp (UMIN R000056536). The requirement for written informed consent was waived due to the study’s retrospective nature. The full study protocol is available to the first author upon request.

**Figure 1.**
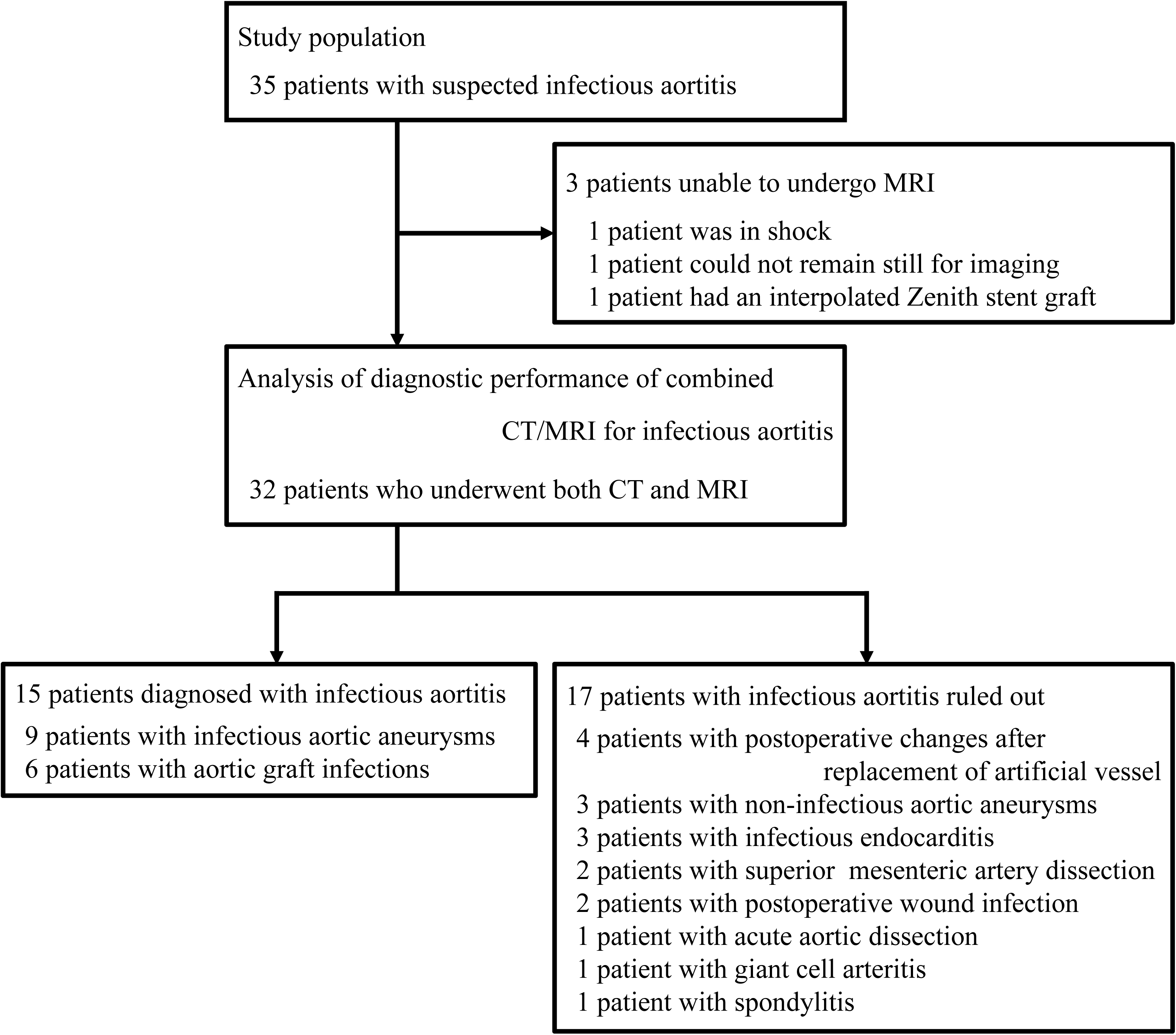
Study flow diagram, including the number of patients with suspected infectious aortitis enrolled and analyzed and the reasons for exclusion. CT, computed tomography; MRI magnetic resonance imaging

### Clinical diagnosis

Clinical diagnosis was obtained from medical records. The diagnosis was usually performed comprehensively with image findings, clinical manifestations (fever, pain, and shock), laboratory findings (elevated inflammatory markers, such as C-reactive protein and white blood cell count), and positive blood or aneurysm wall cultures.^2,21^ Inflammatory findings responsive to antibiotic therapy also supported the diagnosis.^22^

### MRI examinations

MRI was conducted using a 1.5-T scanner (MAGNETOM Avanto, Siemens, Erlangen, Germany). No fasting was required before imaging. Patients were placed in the supine position and immobilized with the body coil in a head-first orientation. The imaging parameters for DWIBS were as follows: field of view (FOV), 450 mm; resolution, 132×132; number of slices, 40; slice thickness, 6 mm; repetition time (TR), 7000 ms; echo time (TE), 66 ms; inversion time (TI), 180 ms; number averages, 5; b-value, 800 s/mm^2^; free-breathing; mode, parallel imaging technique (PAT) for generalized autocalibrating partially parallel acquisitions (GRAPPA2); and total scan duration, 2 min and 41 s. For T2-weighted imaging with a half-fourier acquisition single-shot turbo spin-echo (HASTE) sequence, the imaging parameters were as follows: FOV, 450 mm; resolution, 256×154; the number of slices, 40; slice thickness, 6 mm; TR, 600 ms; TE, 83 ms; breath-holding; and scan duration, 35 s. For T1-weighted imaging with a volumetric interpolated breath-hold examination (VIBE) sequence, the imaging parameters were as follows: FOV, 450 mm; resolution, 320×168; number of slices, 80; slice thickness, 3 mm; TR, 6.91 ms; and TE, 2.39 ms. No contrast agents were administered during imaging.

### CT examinations

A 256-multislice CT (MSCT) system (SOMATOM drive, Siemens, Erlangen, Germany) or 128-MSCT system (SOMATOM Definition Flash, Siemens) was used for CT examinations. Contrast agents were used in some cases, but this study employed non-contrast CT images. The imaging parameters were as follows: voltage, 100–120 kV; slice thickness, 1 mm; pitch factor, 1.2; and FOV, 300–400 mm.

### Image diagnosis

Image diagnosis was performed for each of CT, DWIBS, and combination of CT and DWIBS (combined DWIBS/CT) by two experienced radiologists blinded to clinical information using a SYNAPSE VINCENT workstation (Fujifilm Corporation, Tokyo, Japan). A positive diagnosis was established for combined DWIBS/CT only when both DWIBS and CT were positive. If the two readers’ opinions differ, they discuss them together for a final qualitative evaluation.^10,23^

The findings for a positive CT diagnosis were a periaortic mass, periaortic gas, rapid aortic dilation, and a change in aortic morphology over time.^2^ Moreover, specific findings associated with aortic graft infection included swelling around the prosthetic graft, the formation of a fistula or pseudoaneurysm, and the retention of fluid or gas around the graft.^21^ DWIBS was positive when a higher signal area was around the artery than the reference object.^24–26^ The signal intensity on the spinal cord (spinal cord) (DWIBS [spinal cord]) and the normal aorta (DWIBS [aorta]) was used as reference. The apparent diffusion coefficient (ADC) values were not evaluated because the abscess lumen had a non-circular morphology with heterogeneous luminal components, and there were no high-signal areas when the abscess lumen was not present.^27^

### Statistical analysis

The analyses conducted in this study were pre-specified, based on a protocol defined strictly before data collection commenced. Continuous variables were described as medians (interquartile range [IQR]). Categorical variables were represented as frequencies and percentages. Sensitivity, specificity, and positive and negative predictive values are presented with corresponding 95% confidence intervals (CIs). Receiver operating characteristic (ROC) curves and area under the curve (AUC) were calculated. We also calculated effect estimates with their 95% CIs to assess the diagnostic parameters and performed DeLong’s tests for each ROC curve. We then assessed the inter-rater reliability between the two readers using Cohen’s kappa coefficient. The logistic regression analysis and decision curve analysis (DCA) were performed to assess the benefit of adding DWIBS to CT. A logistic regression model was constructed using DWIBS and CT as independent variables. Then, the AUC was calculated for the model, and the difference between the DWIBS+CT model and the CT alone model was compared using DeLong’s test. DCA analysis was conducted to examine whether the DWIBS+CT model is superior to the CT alone model at a certain range of threshold probability with respect to the net benefit.

In the DCA analysis, a net benefit of clinical judgment to opt or not opt for the treatment based on a prediction model was assessed by comparing harm (treating a patient without disease) to benefit (treating a patient with disease). The harm: benefit ratio corresponds to a threshold probability where the expected benefit of treatment equals the expected benefit of avoiding treatment.^28^ In the opt-in policy, the net benefit of receiving treatment was calculated using true positive and false positive, resulting in the higher net benefit in the setting of the smaller harm: benefit ratio. Conversely, in the opt-out policy, the net benefit of avoiding treatment was obtained from true negative and false negative and was higher if the harm: benefit ratio became greater. Net benefits were graphically represented against threshold probabilities to discern the superior model in terms of clinical utility.^28–30^

Statistical analyses were performed using R (version 4.2.2; R Foundation for Statistical Computing, Vienna, Austria), with the "rmda" package utilized for DCA. P-values ≤ 0.05 were considered statistically significant (two-tailed). There were no missing values in the dataset, therefore, handling of missing data was not considered in this study.

## Results

### Characteristics of the patients

Table 1 presents the characteristics of the patients. The median age was 76 years (IQR, 46–94 years), and 9 (28%) were female. Fifteen (47%) patients were diagnosed with infectious aortitis, including 9 (28%) with infectious aortic aneurysm and 6 (40%) with aortic graft infection. The patients diagnosed with infectious aortitis had fever in 15 (100%), pain in 10 (67%), and positive blood culture in 11 (73%) patients. The diagnosis of 17 (53%) patients without infectious aortitis was non-infectious aortic aneurysms in three, acute aortic dissections in two, infectious endocarditis in three, superior mesenteric artery dissections in two, postoperative wound infections in two, giant cell arteritis in one, and spondylitis in one patient (Table S1).

**Table 1.** Baseline characteristics of the patients with suspected infectious aortitis.

| Characteristic | Overall<br>N = 32 | Infectious Aortitis<br>N = 15 | Non-Infectious<br>Aortitis<br>N = 17 |
| --- | --- | --- | --- |
| Age, years<sup>*</sup> | 76 (46, 94) | 81 (59, 92) | 70 (46, 94) |
| Male sex, n | 23 (72%) | 9 (60%) | 14 (82%) |
| Height, cm<sup>*</sup> | 165 (142, 174) | 162 (142, 174) | 166 (150, 173) |
| Weight, kg<sup>*</sup> | 55 (31.5, 79.2) | 50 (39.5, 79.2) | 58 (31.5, 76.7) |
| Body mass index, kg/m<sup>2*</sup> | 20.8 (14.0, 27.5) | 19.6 (15.9, 27.5) | 21.0 (14.0, 26.6) |
| Pain, n | 19 (59%) | 10 (67%) | 9 (53%) |
| Fever, n | 23 (72%) | 15 (100%) | 8 (47%) |
| Shock, n | 2 (6.2%) | 1 (6.7%) | 1 (5.9%) |
| Culture positive, n | 17 (53%) | 10 (67%) | 7 (41%) |
| C-reactive protein<sup>*</sup> | 14.8 (0.04, 26.5) | 17.3 (8.30, 26.3) | 10.4 (0.04, 26.5) |
| Leukocyte count, 10<sup>3</sup>/μL<sup>*</sup> | 9.80 (3.00, 3.18) | 12.1 (8.30, 18.1) | 6.40 (3.00, 31.8) |
| Procalcitonin, ng/mL<sup>*</sup> | 0.27 (0.02, 21.0) | 0.51 (0.04, 5.62) | 0.12 (0.02, 21.0) |
| Ascending, n | 3 (9.4%) | 0 (0%) | 3 (18%) |
| Arch, n | 15 (47%) | 9 (60%) | 6 (35%) |
| Descending, n | 5 (16%) | 3 (20%) | 2 (12%) |
| Paravisceral, n | 4 (12%) | 1 (6.7%) | 3 (18%) |
| Infrarenal, n | 4 (12%) | 2 (13%) | 2 (12%) |
| Iliac, n | 2 (6.2%) | 2 (13%) | 0 (0%) |
| Multiple, n | 4 (12%) | 3 (20%) | 1 (5.9%) |
| Hypertension, n | 15 (47%) | 9 (60%) | 6 (35%) |
| Diabetes mellitus, n | 5 (16%) | 4 (27%) | 1 (5.9%) |
| Lipid disorders, n | 8 (25%) | 4 (27%) | 4 (24%) |
| Chronic kidney disorder, n | 6 (19%) | 3 (20%) | 3 (18%) |
| Cancer, n | 3 (9.4%) | 2 (13%) | 1 (5.9%) |
| Post-aortic surgery, n | 17 (53%) | 6 (40%) | 11 (65%) |
| Within 3 months, n | 5 (16%) | 0 (0%) | 5 (29%) |
| Open surgery, n | 9 (28%) | 5 (33%) | 4 (24%) |
| Endovascular surgery, n | 4 (12%) | 4 (27%) | 1 (5.9%) |
| n (%); * Median (IQR) |  |  |  |

Among patients diagnosed with infectious aortitis, eight (53%) patients underwent surgical treatment, including prosthetic graft replacement and stent grafting in five (33%) and four (27%) patients, respectively. One patient underwent stent grafting followed by prosthetic graft replacement as a planned two-stage surgery. One patient required an unplanned second open surgery after stent grafting due to reinfection. Another patient died from sepsis due to reinfection after stent grafting. The other six patients had no major complications. In contrast, seven (47%) patients who did not undergo surgery, including stent grafting, were treated with antibiotics owing to poor condition or patient refusal. Among them, four patients died from events related to aortic infection: sepsis in three and aortic rupture in one patient.

No MRI (including DWIBS)-associated adverse events, such as dizziness or nausea after the test, were observed.

### Diagnostic performance of DWIBS and CT

Table 2 presents the diagnostic performance. DWIBS was positive in 18 (56%) and 23 (72%) patients with the spinal cord and aorta as a reference, respectively. The sensitivity, specificity, and AUC of DWIBS (spinal cord) were 93.3%, 76.5%, and 0.85, respectively. In contrast, those for DWIBS (aorta) were 93.3%, 47.1%, and 0.70, respectively. The inter-reader kappa coefficients of DWIBS (spinal cord) and DWIBS (aorta) were 0.875 and 0.855, respectively. CT was positive in 19 patients. The sensitivity, specificity, AUC, and inter-rater kappa coefficient of positive CT diagnosis were 93.3%, 70.6%, 0.82, and 0.529, respectively (Table S2).

**Table 2.** Diagnostic performance of each imaging modality.

|  | TP | FP | TN | FN | Sensitivity | Specificity | PPV | NPV | AUC |
| --- | --- | --- | --- | --- | --- | --- | --- | --- | --- |
| DWIBS<br>(aorta)* | 14 | 9 | 8 | 1 | 93.3<br>(68.1–99.8) | 47.1<br>(23.0–72.2) | 60.9<br>(38.5–80.3) | 88.9<br>(51.8–99.7) | 70.2<br>(56.3–84.1) |
| DWIBS<br>(spinal cord)† | 14 | 4 | 13 | 1 | 93.3<br>(68.1–99.8) | 76.5<br>(50.1–93.2) | 77.8<br>(52.4–93.6) | 92.9<br>(66.1–99.8) | 84.9<br>(72.6–97.2) |
| CT | 14 | 5 | 12 | 1 | 93.3<br>(68.1–99.8) | 70.6<br>(44.0–89.7) | 73.7<br>(48.8–90.9) | 92.3<br>(64.0–99.8) | 82.0<br>(69.0–94.9) |
| DWIBS+CT<br>(aorta)* | 13 | 5 | 12 | 2 | 86.7<br>(59.5–98.3) | 70.6<br>(44.0–89.7) | 72.2<br>(46.5–90.3) | 85.7<br>(57.2–98.2) | 78.6<br>(64.4–92.9) |
| DWIBS+CT<br>(spinal cord)† | 13 | 1 | 16 | 2 | 86.7<br>(59.5–98.3) | 94.1<br>(71.3–99.9) | 92.9<br>(66.1–99.8) | 88.9<br>(65.3–98.6) | 90.4<br>(79.8–100.0) |

n, % (% confidence interval); Abbreviations: TP, true positive; FP, false positive; TN, true negative;
TN, true negative; PPV, positive predictive value; NPV, negative predictive value; AUC, area under the curve;
DWIBS, diffusion-weighted whole-body imaging with background suppression; CT, computed tomography
\*aorta was used as a reference.
†spinal cord was used as a reference.

In contrast, DWIBS+CT was positive in 14 (44%) and 18 (56%) patients for DWIBS (spinal cord) and DWIBS (aorta), respectively. The sensitivity, specificity, and AUC of DWIBS (spinal cord) +CT were 86.7%, 94.1%, and 0.90, respectively. In contrast, for DWIBS (aorta) +CT, they were 86.7%, 70.6%, and 0.79, respectively. The AUC for DWIBS (spinal cord) +CT was significantly higher than that for DWIBS (aorta) +CT (difference 11.8%; 95% CI: 1.37%–22.2%; P=0.027).

### Diagnostic model

The logistic regression model for the prediction of aortic infection was constructed using DWIBS and CT as independent variables. The regression coefficients for DWIBS and CT were 4.07 (95% CI: 1.17–6.97) and 3.78 (95% CI: 0.84–6.72), respectively. The equation to calculate the probability of aortic infection is given below:

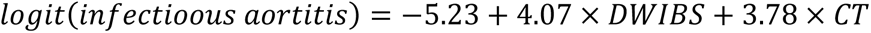

where DWIBS or CT = 1 if each examination was performed.

The AUC for DWIBS (spinal cord)+CT was 0.94 (95% CI: 0.86–0.10), which tended to be higher than the AUC for CT alone (difference 12.1%, 95% CI: -24.6%–0.290%; P= 0.0557) (Figure 2A).

**Figure 2.**
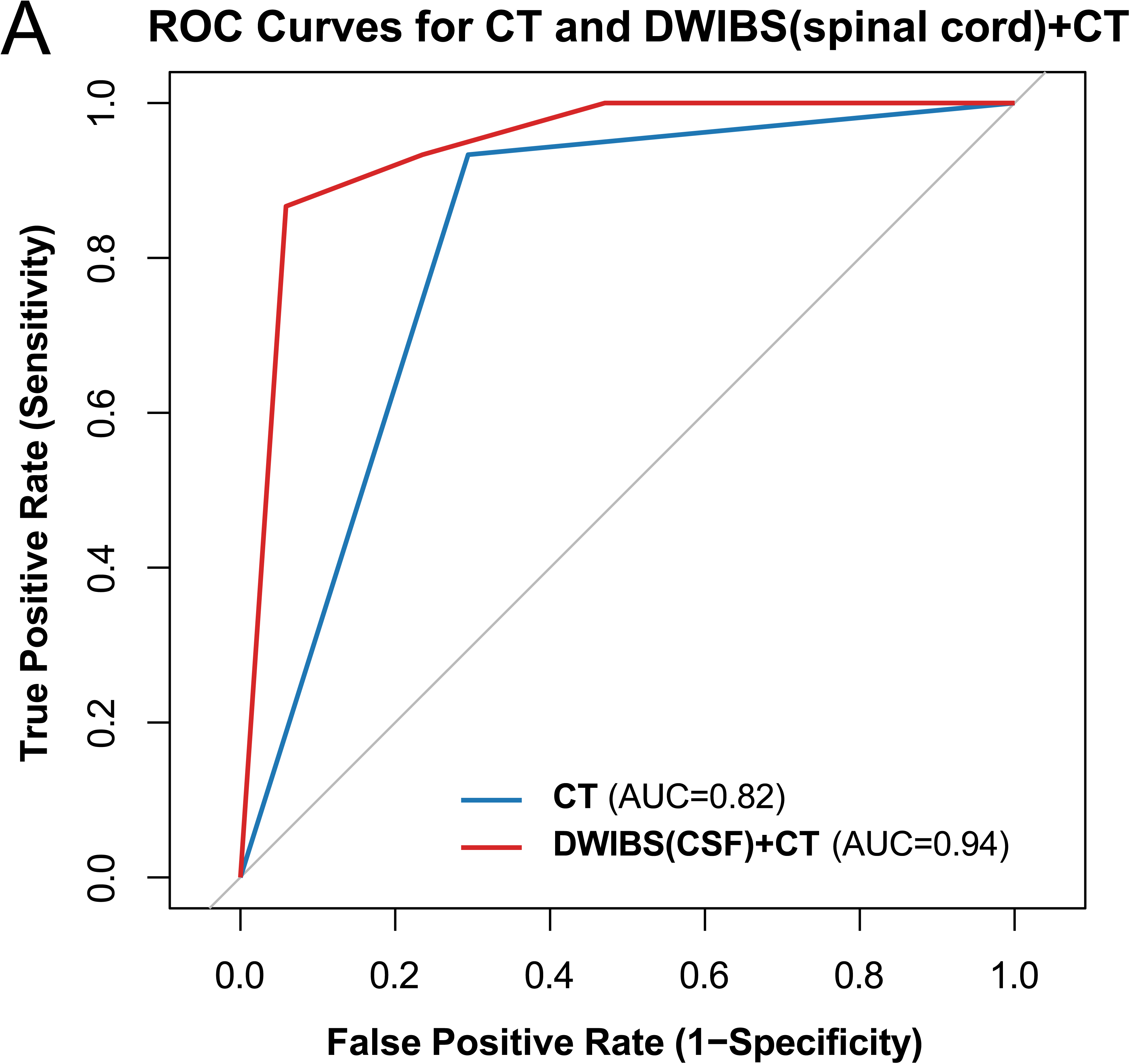
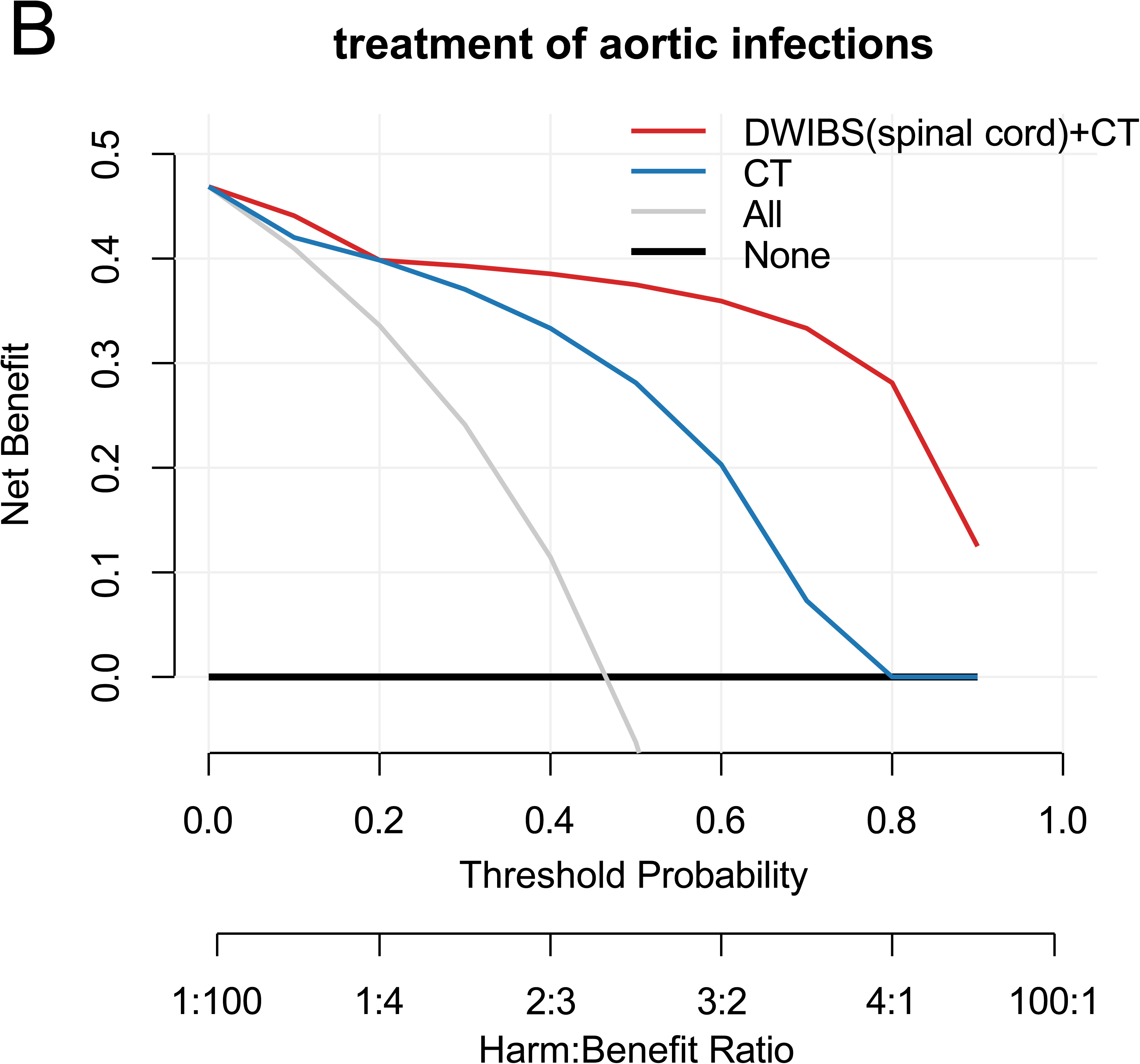
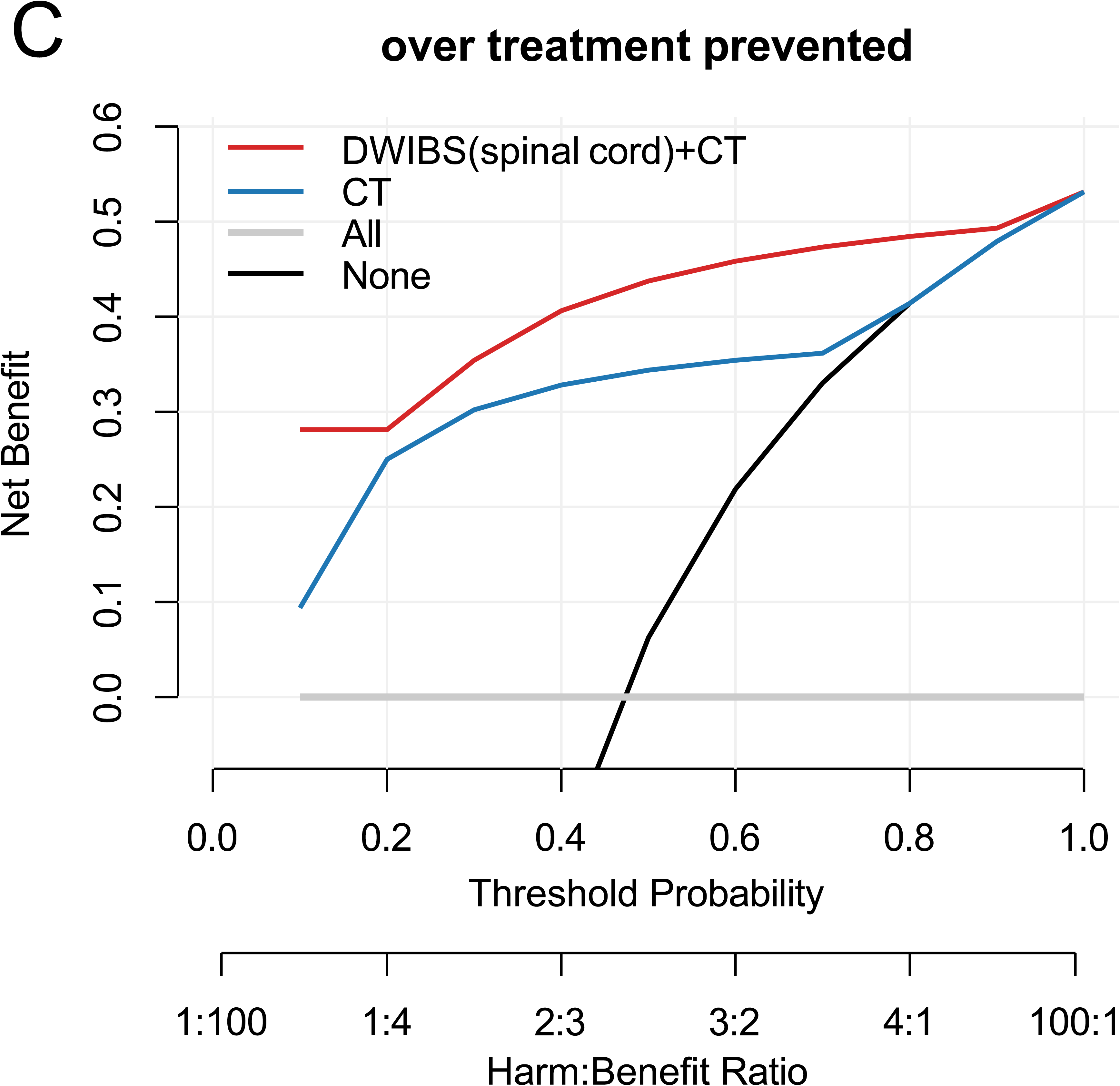
Comparison of diagnostic performance between DWIBS and CT. (A) ROC curves for predicting infectious aortitis using CT alone and the combination of DWIBS (spinal cord) and CT. (B) Net benefits of receiving treatment of infectious aortitis based on CT-only model and DWIBS+CT model. (C) Net benefits of avoiding treatment based on each model. Net benefits when all patients received or avoided treatment were also given.

The DCA analysis demonstrated that the DWIBS+CT model had superior net benefit to the CT alone model on almost all threshold probabilities in both opt-in and opt-out policies. The net benefit of receiving treatment was higher in the DWIBS+CT model than in other models, especially at a threshold probability of greater than 20% (Figure 2B). In contrast, the net benefit of avoiding treatment was higher in the DWIBS+CT model than others throughout all threshold probabilities (Figure 2C).

## Discussion

To our knowledge, this is the first study to investigate the diagnostic performance of DWIBS for infectious aortitis. This study demonstrated that combined DWIBS (spinal cord)/CT had a higher diagnostic performance than CT.

PET/CT is considered the most reliable modality for diagnosis of infectious aortic aneurysms with reported sensitivity, specificity, and AUC of 100% (95% CI: 75.3%–100%), 57.1% (95% CI: 34.0%–78.2%), and 0.79 (95% CI: 0.68–0.89), respectively.^19^ For aortic graft infections, combined PET/CT and contrast-enhanced CT had a sensitivity of 100% (95% CI: 75.3%–100%), specificity of 83.3% (95% CI: 35.9%–99.6%), and AUC of 0.93 (95% CI: 0.68–0.10%).^31^ In this study, DWIBS (spinal cord) presented a high diagnostic performance comparable to PET/CT. Furthermore, when combined with CT, sensitivity, specificity, and AUC were 86.7%, 94.1%, and 0.90, respectively, which were comparable to combined PET/CT and contrast-enhanced CT. Considering the feasibility of PET/CT for infectious aortitis, our data suggested that DWIBS can be a viable option for this entity.

The protocols and positive criteria of DWIBS for infectious aortitis should differ from those generally employed for tumor screening. Despite the many quantitative assessments used in cancer diagnosis using ADC values, the non-uniform nature of fluid components in abscess cavities and infected organs flaws the reproducibility of ADC values. Furthermore, although qualitative evaluations are often made by comparing the same tissue, the aorta is luminal and contains few parenchymal components. In addition, the infection can extend over the entire circumference, making it inappropriate to target the same organ. For these reasons, we considered that qualitative evaluation of DWIBS, achieved by comparing spinal cord and signal values, was useful. DWIBS (spinal cord) demonstrated strong reproducibility with minimal inter-reader variability. Therefore, comparison with the spinal cord, which is an organ located within the imaging range and contains fluid components, can be considered a simple and effective approach.

In the present study, the b-value was set to 800 s/mm^2^. While a b-value of 1000 sec/mm^2^ is often used for tumor screening, it was essential to evaluate both the abscess cavity and tissue edema due to infection in the context of aortic infections, necessitating a high-resolution approach.^26^ Higher b-values may reduce the T2 effect, making it difficult to identify tissue edema and potentially increasing the likelihood of false negatives.^32^ Additionally, aortic plaques that are highly fibrotic show low signal values on T2-weighted images; when the infection surrounds a stable plaque, the surrounding signal value is attenuated, and the abscess cavity may be inconspicuous.^33^ Indeed, one false-negative case was observed in DWIBS (spinal cord), underscoring the need for caution in such scenarios. Furthermore, the use of a workstation to add pseudo-colors and combine them with T2-weighted images can simulate images akin to PET/CT, offering an intuitively easier-to-understand diagnostic method (Figure 3).^34^

**Figure 3.**
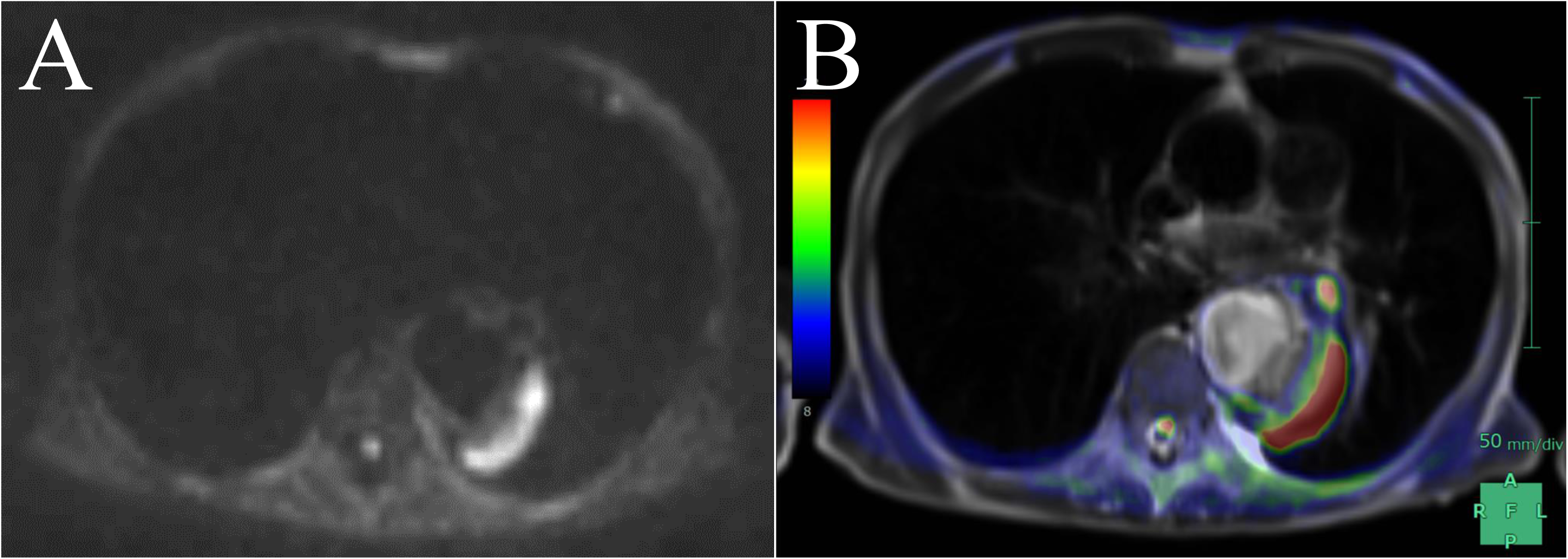
Additive composition of DWIBS and T2-weighted images. (A) On DWIBS imaging, the periaortic abscess shows high signal intensity. (B) A composite of DWIBS and T2-weighted images is shown; the combination of DWIBS with pseudo-colored T2-weighted images produces a composite image resembling PET/CT, providing enhanced visualization and accurate localization of infectious spill-over and associated anatomical structures. DWIBS, diffusion-weighted whole-body imaging with background suppression; CT, computed tomography; PET, positron emission tomography

Infectious aortitis is associated with relatively greater harm of treating patients without disease because the radical treatment of this entity is surgical and usually highly invasive. Therefore, the DCA analysis is quite effective because it provides a net benefit of clinical judgment based on each diagnostic modality over a range of harm-to-benefit ratios. In this study, the DCA analysis supported the efficacy of adding DWIBS to non-contrast CT to diagnose infectious aortitis. In an opt-in approach, a certain level of net benefit was obtained with non-contrast CT alone at a low threshold probability; however, a higher net benefit was obtained by adding DWIBS, especially in cases with higher harm: benefit ratio. This suggested that diagnosis using CT alone would be sufficiently beneficial to initiate less invasive treatment, such as medication with antibiotics, whereas adding DWIBS would be more effective for decision-making to opt for more invasive treatment. In contrast, in an opt-out approach, DWIBS with CT also showed a greater net benefit of avoiding treatment than that of CT alone. However, the difference between them was relatively constant throughout all threshold probabilities. These findings suggested that adding DWIBS to CT was beneficial in the clinical judgment of treatment for aortic infections, especially in adopting invasive treatments.

This study had some limitations. Firstly, this was a retrospective, single-center study; selection bias cannot be excluded. Secondly, while our study emphasizes the diagnostic utility of combining DWIBS and CT for infectious aortitis, there are inherent limitations in our diagnostic model. The model is formed solely based on CT and DWIBS, raising concerns about the potential for overfitting. Despite the insights provided, it’s paramount to view the diagnostic model analysis as supplementary information and exercise caution when interpreting its implications. Nonetheless, the criteria for suspicion were clearly defined, and consecutive cases were included in the study. In addition, we consider racial disparities unlikely to affect the diagnosis of infectious aortitis. Furthermore, because qualitative assessment is performed, discerning aortitis or inflammatory aortic aneurysms from an acute thrombosed aortic dissection, which also exhibits high signal values on DWI, may be difficult and false-positive results may arise when DWIBS is performed in the early postoperative period after surgical interventions such as prosthetic graft replacement because the signal is high for fluid components. A comprehensive evaluation using contrast-enhanced CT and serum markers is necessary (Figure 4).

**Figure 4.**
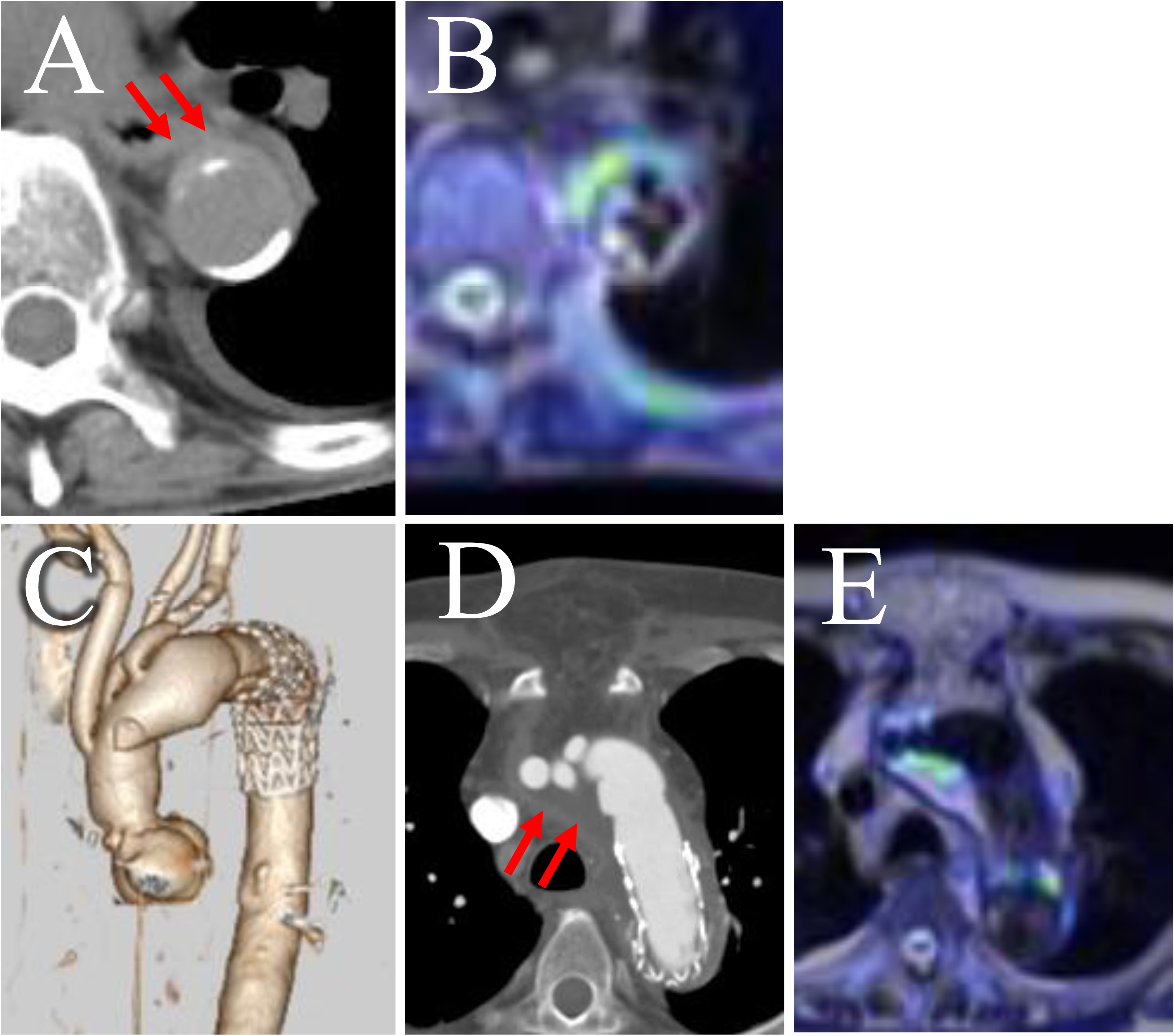
Pitfalls of DWIBS. Integration of multiple imaging modalities is essential for an accurate diagnosis, as MRI, especially DWIBS and T2-weighted images, alone may be inadequate for comprehensive evaluation. (A–B) Depiction of thrombus occlusion-type aortic dissection, where non-contrast CT reveals false lumen CT values while DWIBS signals reflect acute hematoma (arrows). (C–E) Illustration of an early case of surgery for an infectious aortic aneurysm, wherein caution is warranted due to potential postoperative changes that may confound the DWIBS interpretation (arrows). Despite the partially high signal intensity on DWIBS, no evidence of infection was observed. MRI, magnetic resonance imaging; DWIBS, diffusion-weighted whole-body imaging with background suppression; CT, computed tomography

The results of this study suggest that the combination of CT and DWIBS, a previously underutilized MRI technique, offers a promising diagnostic approach for patients with suspected infectious aortitis.

## Data Availability

The data supporting the findings of this study are available from the corresponding author, JS, upon reasonable request.

## Non-standard Abbreviations and Acronyms

DWIBS: diffusion-weighted whole-body imaging with background body signal suppression;
FOV: field of view;
TR: repetition time;
TE: echo time;
TI: inversion time;
PAT: parallel imaging technique;
GRAPPA2: generalized autocalibrating partially parallel acquisitions;
HASTE: half-Fourier acquisition single-shot turbo spin-echo;
VIBE: volumetric interpolated breath-hold examination;
MSCT: multislice computed tomography;
ADC: apparent diffusion coefficient;
DCA: decision curve analysis.

## Acknowledgments

We want to thank Editage (www.editage.com) for English language editing.

## Supplementary Material

Tables S1 and S2

## Author contributors

JS contributed to the study design, interpretation of the results, and manuscript writing. MM contributed to the study design, planning, and manuscript review. MT conducted the data analysis and revised the manuscript. SK, NY, and HS collected the data and critically reviewed the manuscript. TH and MA assisted with interpreting the results and reviewed the manuscript. SW summarized the study and revised the manuscript. All authors approved the final version of the manuscript.

